# Predicting disability progression and cognitive worsening in multiple sclerosis using grey matter network measures

**DOI:** 10.1101/2021.03.23.21253388

**Authors:** Elisa Colato, Jonathan Stutters, Carmen Tur, Narayanan Sridar, Douglas Arnold, Claudia Wheeler Kingshott, Frederik Barkhof, Olga Ciccarelli, Declan Chard, Arman Eshaghi

**Author notes:** **Corresponding Author:** Arman Eshaghi, Queen Square Multiple Sclerosis Centre, Department of Neuroinflammation, UCL Queen Square Institute of Neurology, University College London, London, UK.

## Abstract

**Objective:** In multiple sclerosis (MS), magnetic resonance imaging (MRI) measures at the whole brain or regional level are only modestly associated with disability, while network-based measures are emerging as promising prognostic markers. We sought to demonstrate whether data-driven network-based measures of regional grey matter (GM) volumes predict future disability in secondary progressive MS (SPMS).

**Methods:** 

We used cross-sectional structural MRI, and baseline and longitudinal data of Expanded Disability Status Scale [EDSS], 9-Hole Peg Test [9HPT], and Symbol Digit Modalities Test [SDMT], from a clinical trial in 988 people with progressive MS. We processed T1-weighted scans to obtain GM probability maps and applied spatial independent component analysis (ICA) to identify co-varying patterns of GM volume change. We used survival models to determine whether baseline GM network measures predict cognitive and motor worsening.

**Results:** We identified 15 networks of regionally co-varying GM features. Compared with whole brain GM, deep GM, and lesion volumes, ICA-components correlated more closely with clinical outcomes. A mainly basal ganglia component had the highest correlations at baseline with the SDMT and was associated with cognitive worsening (HR= 1.29, 95% CI [1.09-1.52], p< 0.005). Two ICA-components were associated with 9HPT worsening (HR=1.30, 95% CI [1.06:1.60], p<0.01; and HR= 1.21, 95%CI [1.01:1.45], p<0.05). Post-hoc analyses revealed that for 9HPT and SDMT survival models including network-based measures reported a higher discrimination power (respectively, C-index= 0.69, se= 0.03; C-index= 0.71, se= 0.02) compared to models including only whole and regional MRI measures (respectively, C-index= 0.65, se= 0.03; C-index= 0.69, se= 0.02).

**Conclusions:** The disability progression was better predicted by networks of covarying GM regions, rather than by single regional or whole-brain measures. Network analysis can be applied in future clinical trials and may play a role in stratifying participants who have the most potential to show a treatment effect.

## Introduction

Multiple sclerosis (MS) is an inflammatory and neurodegenerative disease of the central nervous system (CNS). The most recognised pathological feature of MS is an inflammatory demyelinating white matter (WM) lesion, whose formation is associated with relapses. However, the principal driver of irreversible disability, and progressive MS, is thought to be neurodegeneration [1,2]. We now have many treatments that reduce the risk of MS relapses, but only have two licensed treatments for progressive MS, and their efficacy appears to be mainly in people who still show evidence of ongoing inflammatory lesion activity.

Neurodegeneration manifests as brain atrophy and this can be measured with magnetic resonance imaging (MRI) [3]. Brain atrophy is mainly due to GM volume loss, GM volume loss is faster in deep grey matter (DGM) than the cortex, and within the cortex preferentially affects temporal and parietal regions [2,4,5]. In phase 2 progressive MS clinical trials MRI-based measures of brain atrophy are now the preferred outcome, as they have proven more sensitive to treatment effects than clinical measures [6,7]. However, regional and global brain atrophy, and other conventional MRI measures, only partly correlate with and predict disability progression in people with progressive MS [8]. In part, this is explained by pathology being assessed at a whole or regional brain level, while the disability occurs as a result of impaired connections between clinically eloquent regions.

Pathology in MS affects some parts of the brain more than others, and ideally, we should seek to measure pathology where it is most likely to affect clinical outcomes. With this in mind, network-based measures have the potential to add value to conventional MRI measures, and have already proven promising in explaining motor disability [9]. Data-driven GM network measures are also a good candidate to be used as prognostic markers in clinical trials, and these are important as they can potentially substantially enrich clinical trials with patients more likely to progress and so demonstrate treatment effects.

Independent component analysis (ICA) is a robust data-driven technique that has been used to identify brain networks on structural MRI [10,11]. Spatial ICA can identify separate brain regions whose volume covaries, which can be linked by a common biological or pathological property [12,13]. In a mainly relapsing-remitting (RR) MS cohort, in a cross-sectional study Steenwijk and colleagues (2016) used the associated technique of canonical correlations to identify co-varying patterns in cortical thickness associated with clinical outcomes. A previous study in early RRMS showed that co-varying patterns of GM intensities at baseline did not predict confirmed disability progression (CDP) within 10 years, or at 10 years differentiate between patients with CDP and without CDP [14]. These studies were weighted towards RRMS, and while atrophy occurs early in MS, it is more prominent, and thought to be more clinically relevant [15,16], during the progressive phase. No study so far has looked at the predictive value of baseline network-based measures of the cortex and DGM.

The overarching goal of our study was to apply network-based MRI measures of GM atrophy, seeking to better predict disability progression in secondary progressive MS when compared to conventional regional or whole brain volumetric measures. We applied spatial ICA to identify co-varying patterns of GM from structural MRI in 988 people with SPMS. Our specific aims were to (1) identify clinically relevant GM network measures at study entry, and (2) identify GM network measures that predict future disability progression. We also aimed to assess stability and reliability of these networks.

## Materials and methods

### Participants

We re-analysed data from the ASCEND trial, an international (163 sites across 17 countries), phase 3, randomised, double-blind, placebo-controlled trial. Baseline and longitudinal clinical and baseline MRI data from 1003 subjects aged 18 to 58 years old, who had secondary progressive multiple sclerosis (SPMS), and baseline EDSS score between 3.0-6.5, were acquired [17]. We included visits that acquired the following MRI sequences: (1) T2-FLAIR, (2) T2-weighted, and (3) T1-weighted without contrast administration MRI scans. We excluded data from (n=15) participants with artefacts on the available scans (e.g. ghosting, magnetic susceptibility, and motion artefacts).

## MRI acquisition and processing

### Image acquisition

Brain scans were acquired with 2D T1-weighted with voxel size= 0.98×0.98×3mm^3^; (2) fast fluid-attenuated inversion recovery (FLAIR) with voxel size= 0.98×0.98×3mm^3^, and (3) T2-weighted sequences with voxel size= 0.98×0.98×3mm^3^. Details on MRI acquisition from a representative centre are provided in the *Supplementary Materials*.

### Image Processing

The aim of image processing was to extract GM probability maps which are the input to ICA analysis from T1-weighted MRI. We followed the steps shown in (***Figure 1***).

**Figure 1.**
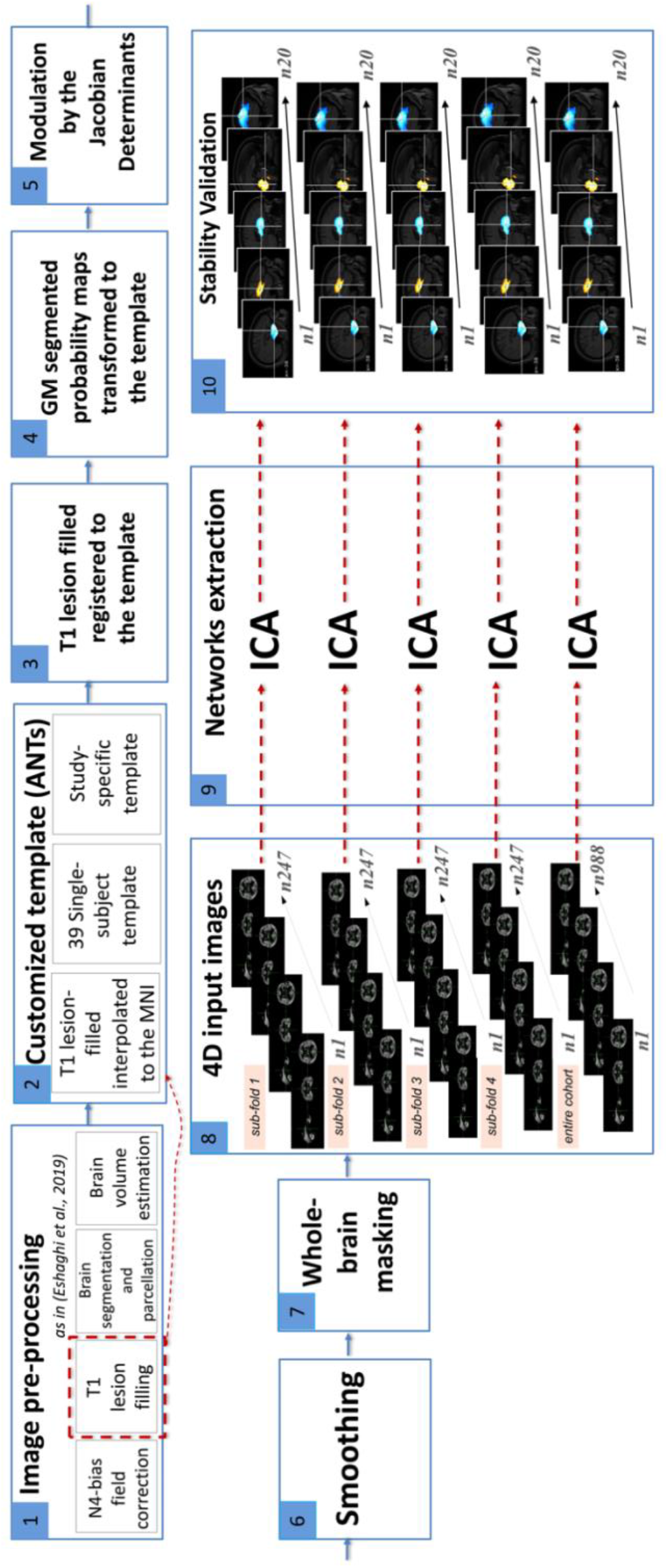
Visual representation of our image-analysis pipeline. Figure legend: Aiming to identify data-driven network-based measures of covarying GM volumes, we initially pre-processed our data as in *Eshaghi et al*., *2019* (N4 bias field correction, lesion filling, brain segmentation and parcellation). We created a customised template from all the available scans from 39 randomly selected subjects. After having resampled those scans to an isotropic space, we created 39 single subject templates, and from those an average study-specific template. We registered the T1 lesion filled scans to the template and diffeomorphically transformed the GM segmentation maps to the template using the warping matrix generated from the previous step. We modulated the GM segmentation maps by the Jacobian determinants in order to account for possible deformations to the original volumes occurred after the non-linear transformation. We applied an 8mm smoothing kernel to account for inter-subject variability and applied a whole brain mask to constrain the following analysis at the level of the brain. Aiming to prove the stability of our results, we randomly divided our cohort into 4 sub-folds. For each sub-fold and for the entire cohort, we generated a 4D image by concatenating the available GM maps and ran fastICA on each of those inputs allowing for 20 components to be identified. For each fold and for the entire cohort, we generated a 4D image by concatenating the 20 generated ICA-components and ran cross-sectional correlations between those inputs to identify which components were stable and could be implemented for statistical analysis. Acronyms: ANTs= Advanced normalization tools; ICA= independent component analysis;

We used an established pipeline as described elsewhere [18]. Briefly, this pipeline included N4 bias field correction [19], lesion filling [20] (to reduce the effects of hypointense lesions in T1 scans during segmentation), and used the Geodesic Information Flows (GIF) version 3.0 [21] to segment the lesion filled T1-weighted images into GM, white matter (WM), and CSF probability maps, as well as to parcellate the brain into 120 regions according to the Neuromorphometrics atlas [22]. We used GIF because it allows the inclusion of 2D-MRI data and does not require additional manual editing, which for a cohort of this size would have been unfeasible. We (EC, AE, DC) visually inspected these outputs to check for erroneous segmentation or parcellation outputs.

### Voxel based analysis of GM probability maps

We randomly selected 39 participants to create a study-specific template (as described in *Supplementary Materials*), to improve the accuracy of the registration and minimise bias [23]. For each participant, we registered the baseline T1-lesion filled scans to the study-specific template using rigid, affine and diffeomorphic non-linear registrations [24]. We calculated the cortical and deep GM volumes from GM probability maps in the native space. We transformed GM probability maps to the template by applying the warping matrices obtained from the previous step. We modulated the GM probabilistic maps by the Jacobian determinants estimated in Advanced Normalization tools (ANTs) version 2.3.1 to adjust for deformations that occurred to the original volumes after the non-linear registration [14,23,25]. We used an 8-mm Full Width at Half Maximum (FWHM) smoothing kernel to account for inter-subject variability. We created a whole brain parcellation mask (as described in *Supplementary Materials*) to constrain the ICA analysis to the brain, and to identify and label brain regions in each ICA-component.

### Network analysis with ICA

We used the FastICA algorithm [26] implemented in scikit-learn 0.23.1 to identify the independent components representing spatial maps of GM co-variation (GM networks). We concatenated the GM probability maps into a 4D volume and fitted the ICA model allowing for 20 components to be identified [11,27]. To assess the stability and repeatability of the identified components, we randomly divided our cohort into 4 folds (247 subjects each) and repeated the analysis for each fold. We generated a 4D image by concatenating the 20 identified components and assessed pairwise spatial cross-correlations with “fslcc” in FSL [28] to select components that were spatially stable for each fold (see ***Figure 1***). We defined components with statistically significant correlations (p<0.05) across sub-folds and entire cohort as stable. We overlaid the stable components with our whole-brain mask (obtained as described in *Supplementary Materials*) to label brain areas involved in each network. To identify a biological meaning, looking at the involved brain areas we compared the identified GM networks with functional networks previously reported in the literature [29,30].

We used the loading factors of the stable components for further statistical analysis. Loading factors quantify the contribution of a given subject to a particular component.

### Statistical analysis

We computed z-scores from the loading factors for each ICA-component, whole brain GM, DGM, and other brain regions volumes with RStudio (version 1.2.5001). To identify components that represent overall brain preservation and brain atrophy, we correlated the loading factors of the ICA-components with baseline whole brain GM volume. We correlated ICA factors with whole brain GM volumes, rather than with the volumes of brain regions involved in each network, because we aimed to determine the direction of ICA-atrophy associations, not their true magnitude (correlations are likely to be smaller than they would otherwise have been considering just brain volumes comprised in each network). To further identify which brain areas in each ICA-component were atrophic and which preserved, we correlated the loading of each network with the baseline volume of the corresponding brain areas. We calculated Pearson correlation coefficients between z-scores of ICA-components and baseline average (dominant and non-dominant hands) 9HPT and SDMT, and Spearman correlations between these z-scores and the baseline EDSS. We calculated correlation coefficients for EDSS, 9HPT, and SDMT with the z-scores of lesion load, whole brain GM volume, DGM, and other brain region volumes known to be affected in MS and to be associated with the investigated clinical tests (i.e., thalamus, precuneus, caudate, putamen, pallidum) [31,32]. Correlations were corrected for multiple comparisons using the false discovery rate (FDR; α= 0.05). We used multivariable stepwise regression models to identify the best predictors across 15 networks and whole brain GM volume, deep GM volume, lesion load, age, gender and trial arm as variables of interest. To evaluate the predictive ability of the independent variables while adjusting for centre effects, we fitted a mixed-effect model using the clinical measures (EDSS, 9HPT, and SDMT) as dependent variables, the identified predictors as fixed effect and the centre as random effect.

To calculate time to worsening of physical and cognitive disability we estimated the EDSS progression as an increase of 1 point from a baseline EDSS score of 5.5 or below, or of 0.5 points from baseline EDSS score greater than 5.5, and these scores were confirmed at least at 3 months [7]. We excluded from this estimation all the clinical visits within 30 days of an MS relapse. We also estimated the 9HPT and SDMT worsening as respectively a 20% increase [7,33] and 10% decrease [34,35] with respect to the baseline score.

We performed Cox regression analysis to determine whether the standardised loading of GM networks at baseline could predict the clinical disability. We built one model for each independent variable (i.e., ICA-networks, whole brain atrophy, DGM atrophy, lesion load, and atrophy in smaller regions), adjusting for age, gender, trial arm, and centre, and having the event and the time-to-event as dependent variables.

To determine whether data-driven networks provide added value above regional MRI volumes and lesion loads, we performed post-hoc analysis applying multivariate Cox proportional regression analysis. To identify the best predictive model for 9HPT and SDMT progression, we compared the performance of the three models with the following independent variables:

1. 15 stable ICA-components,
2. 15 ICA-components together with conventionally used MRI measures (whole brain GM, DGM, and lesion load),
3. conventionally used MRI measures

We estimated the concordance index or C-index, which is a measure of the discrimination power of survival models, and represents the proportion of subjects with a progression on the clinical test and a worse outcome predicted by the model (concordant pairs) divided by the total number of possible evaluation pairs [36]. A C-index of 1 represents a perfect model prediction, while a value of 0.5 denotes random prediction. Age, gender, trial arm, and centre were used as covariates for each model.

## Data availability

Processed data and codes used in this study are available upon request from qualified investigators.

## Results

### Participants

For 15 subjects, their scans did not meet our inclusion criteria. Therefore, our final cohort comprised of 988 patients with SPMS (366 men and 622 women with mean age of 46.71±7.70). ***Table 1*** reports the demographic characteristics of these patients.

**Table 1.**
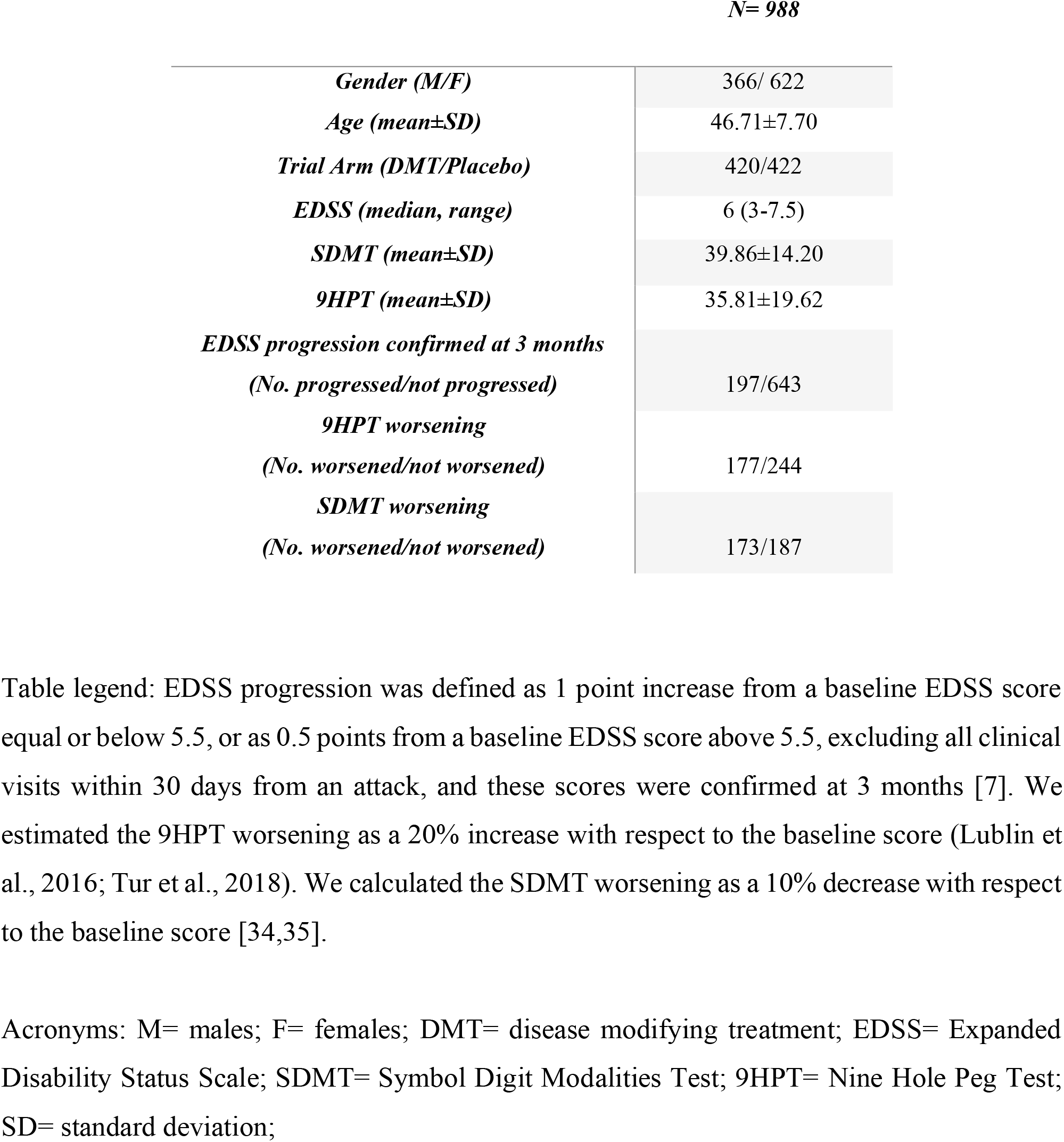
Characteristics of participants

|  | *N= 988* |
| --- | --- |
| *Gender (M/F)* | 366/ 622 |
| *Age (mean±SD)* | 46.71±7.70 |
| *Trial Arm (DMT/Placebo)* | 420/422 |
| *EDSS (median, range)* | 6 (3-7.5) |
| *SDMT (mean±SD)* | 39.86±14.20 |
| *9HPT (mean±SD)* | 35.81±19.62 |
| *EDSS progression confirmed at 3 months*<br>*(No. progressed/not progressed)* | 197/643 |
| *9HPT worsening*<br>*(No. worsened/not worsened)* | 177/244 |
| *SDMT worsening*<br>*(No. worsened/not worsened)* | 173/187 |

Table legend: EDSS progression was defined as 1 point increase from a baseline EDSS score equal or below 5.5, or as 0.5 points from a baseline EDSS score above 5.5, excluding all clinical visits within 30 days from an attack, and these scores were confirmed at 3 months [7]. We estimated the 9HPT worsening as a 20% increase with respect to the baseline score (Lublin et al., 2016; Tur et al., 2018). We calculated the SDMT worsening as a 10% decrease with respect to the baseline score [34,35].
Acronyms: M= males; F= females; DMT= disease modifying treatment; EDSS= Expanded Disability Status Scale; SDMT= Symbol Digit Modalities Test; 9HPT= Nine Hole Peg Test; SD= standard deviation;

### Spatial maps of ICA components overlap with previously known networks

While allowing for up to 20 components, spatial cross-correlation showed that 15 (***Figure 2***) were stable (*Supplementary materials* ***Table 1***).

**Figure 2.**
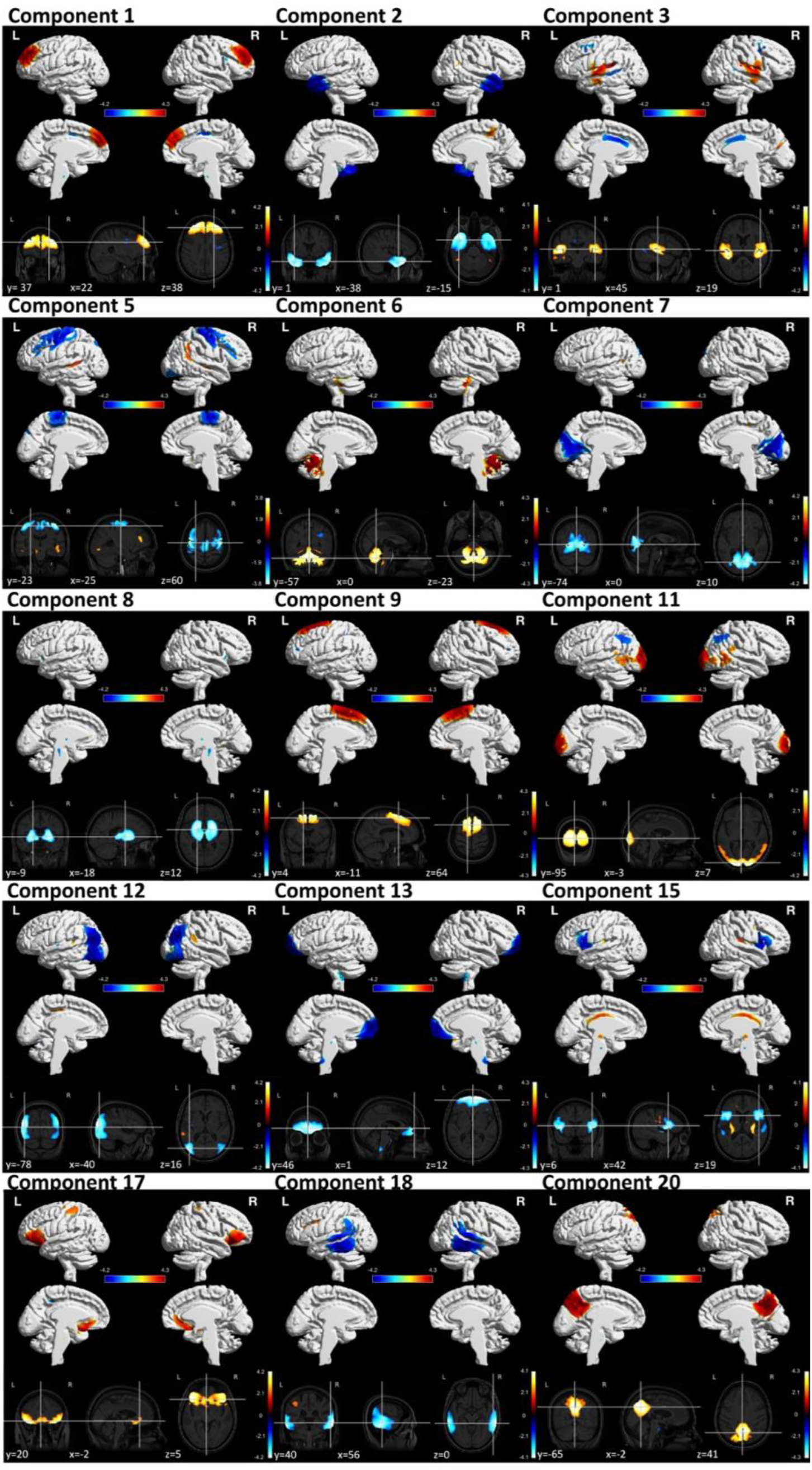
Stable ICA components. Figure legend: To determine the stability of the ICA-networks, we randomly split the sample into four sub-folds and ran the ICA on each of them, as well as on the entire sample. While allowing for 20 components to be identified, cross-sectional correlations proved that only 15 out of the 20 ICA-components were stable (emerged in all of the 4 sub-folds and from the entire sample). The colour bar represents the loading of each component. Most of the identified networks resampled well known functional systems. Component 3 represents an auditory-like network, spanning mainly the superior temporal gyrus, posterior insular and Heschl’s gyrus (cognition-language-speech network). Component 5 is a sensorimotor-like network, encompassing the precentral gyrus, postcentral gyrus, and supramarginal gyrus (action-execution network). Component 6 resamples a cerebellum-like network, involving mainly the cerebellum but also fusiform gyrus, temporal and parietal lobe. Component 8 is a cortico-basal ganglia-like network, spanning the brain stem, pons, thalamus, nucleus accumbens, insula, putamen, caudate, pallidum, frontal and temporal lobe. Component 9 represents an executive control-like network, involving mainly medial frontal areas (action planning and inhibition). Component 11 is a visuo-like network, encompassing mainly several regions of the occipital pole but also supramarginal, temporal and parietal areas. Component 15 resamples a salience-like network, involving the insula, thalamus and striatus (autonomic reaction to salient stimuli; goal directed behaviour). Component 17 represents an affective and reward network, encompassing mainly the anterior cingulate, medial orbitofrontal cortex, and prefrontal cortex. Component 20 resamples a default mode-like network (DMN-like), spanning mainly the precuneus, posterior cingulate, and middle frontal gyrus. The remaining identified networks did not correspond to any major brain functional network, but can be labelled by their predominantly involved brain areas. Component 1 is a superior frontal network, encompassing mainly superior and medial frontal brain areas. Component 2 is a temporal-like network, involving mainly temporal brain regions. Component 7 is a precuneus-like network. Component 12 is an occipito-temporal-like network, spanning mainly the temporal and occipital pole. Component 13 represented a prefrontal cortex-like network, involving mainly frontal and orbitofrontal brain areas. Component 18 is a parieto-temporal-like network, involving mainly temporal and parietal brain areas.

Most of the identified structural GM networks resembled well known functional systems, although the correspondence in the involved regions was never perfect. For example, component 5 is a sensorimotor-like network, encompassing the precentral gyrus, postcentral gyrus, and supramarginal gyrus (action-execution network). Component 8 is a cortico-basal ganglia-like network, spanning the brain stem, pons, thalamus, nucleus accumbens, insula, putamen, caudate, pallidum, frontal and temporal lobe. Component 20 resamples a default mode-like network (DMN-like), spanning mainly the precuneus, posterior cingulate, and middle frontal gyrus. For a detailed description of the remaining networks and of regions associated with each component see ***Figure 2*** and ***Table 2***.

**Table 2.**
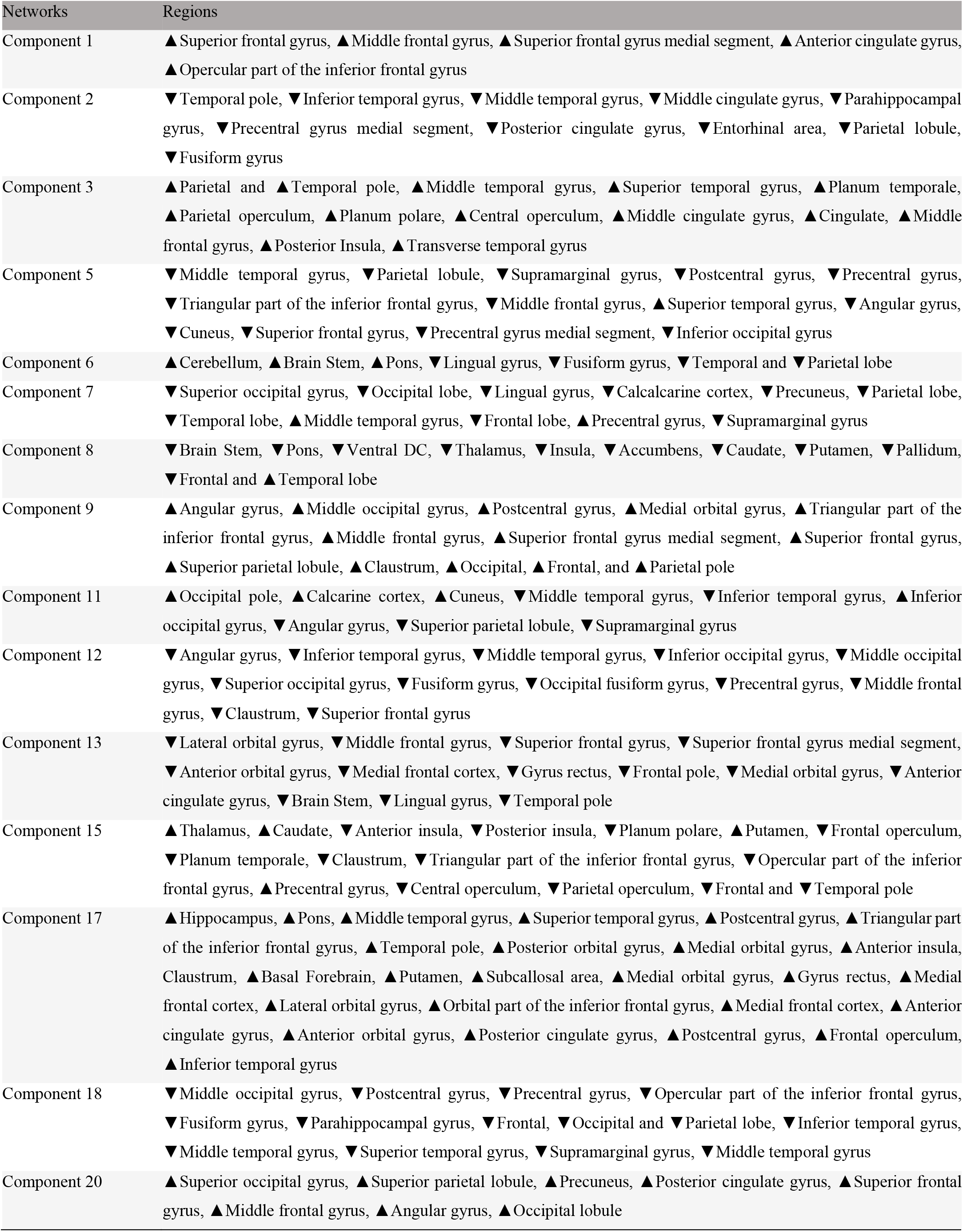
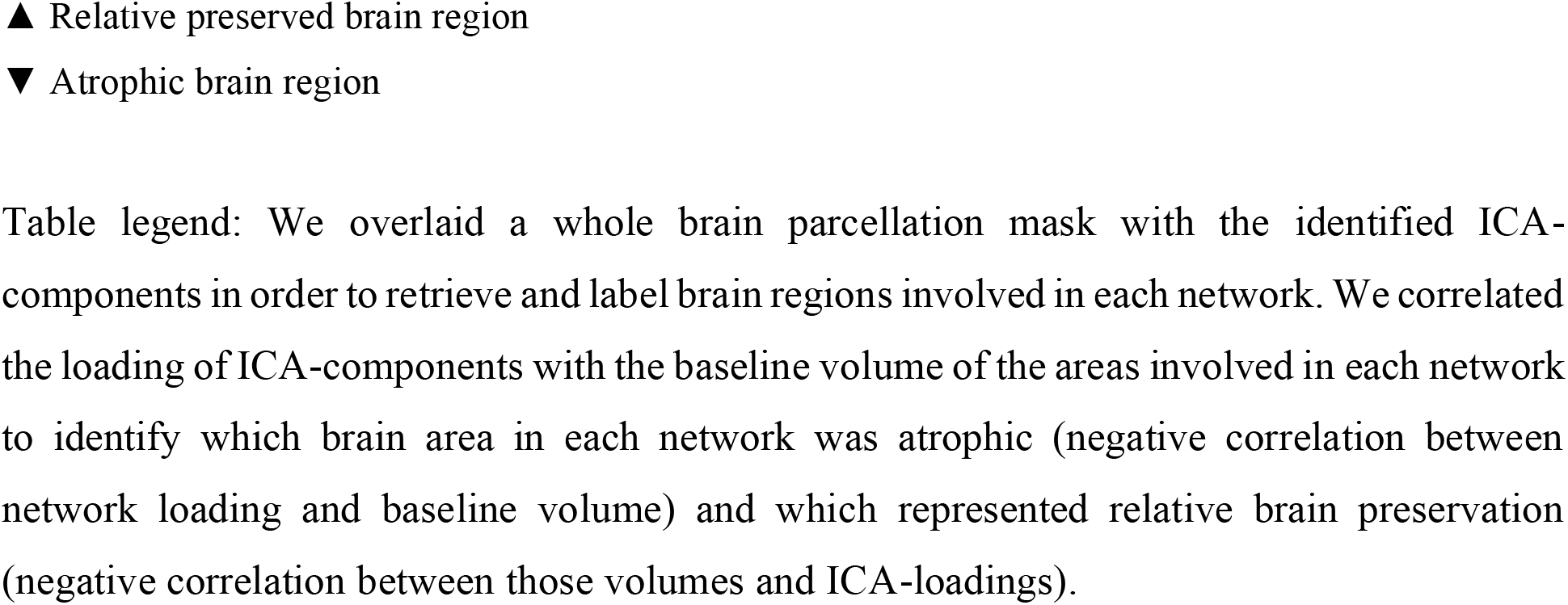
List of the 15 stable components with their corresponding involved brain regions

### Networks represent brain atrophy or preservation

We identified ICA-components representing a mixture of relative brain preservation and brain atrophy. Two representative examples are:

- Component 20 was positively correlated with whole brain GM volumes (r= 0.28, 95% CI [0.22:0.33], p<0.001). Higher component loading was associated with higher GM volume, therefore this component represents a network of relatively greater regional volume at baseline.
- Component 13, instead, was inversely correlated with whole brain GM (r= −0.38, 95% CI [-0.43: −0.33], p<0.001). Higher loading on component 13 was associated with lower whole brain GM volumes, thus this network represents brain atrophy. ***Table 2*** *(Supplementary Materials*) shows correlations between the loading of each ICA-component and whole brain GM volumes.

We identified which brain region in each GM network was atrophic and which preserved (see ***Table 2***).

### Baseline GM networks correlate with clinical measures

Among all ICA-components, component 8 (in which higher values corresponded to lower basal ganglia volumes) was significantly correlated with the SDMT and 9HPT (respectively, r= − 0.44, 95%CI [-0.52: −0.36], p<0.001, and r= 0.18, 95% CI [0.11: 0.24] p<0.001). Component 6 (in which higher values corresponded to higher cerebellar volumes) was correlated with EDSS (rho= −0.11, p= 0.001) (***Figure 3***). Overall, SDMT and 9HPT correlated more strongly with the ICA-components than with conventional MRI measures (***Table 2*** *and* ***Table 3*** *in Supplementary Materials*).

**Table 3.** Comparison between different predictive models for 9HPT and SDMT progression

| Predictors | 20% 9HPT worsening |  |  | 10% SDMT worsening |  |  |
| --- | --- | --- | --- | --- | --- | --- |
|  | Concordance | Standard | Likelihood | Concordance | Standard | Likelihood |
|  | index<br>(C-index) | error (se) | ratio test<br>(p-value) | index<br>(C-index) | error (se) | ratio test<br>(p-value) |
| **15 ICA-components + DGM + whole brain DGM + lesion load** | 0.69 | 0.025 | $p = 9e-05$ | 0.71 | 0.021 | $p = 4e-06$ |
| **15 ICA-components** | 0.68 | 0.025 | $p = 2e-04$ | 0.72 | 0.021 | $p = 5e-06$ |
| **Whole GM + DGM + lesion load** | 0.65 | 0.025 | $p = 8e-05$ | 0.69 | 0.022 | $p = 3e-05$ |

*\*sex, age, trial arm, and centre were used as covariates in each model*
Table legend: Models including ICA-components and conventionally assessed MRI measures, or considering both DGM and the predictive ICA-components identified by Cox-models, have a stronger predictive value when compared to models including just measures of whole brain and deep GM volumes and lesion load. Models including ICA-components and conventionally assessed MRI measures have a stronger predictive value when compared to models including just measures of whole brain and deep GM volumes and lesion load. Concordance index is generally used to validate the predictive ability of survival models. The likelihood ratio test represents the predictive statistically significance value of each model.
Acronyms: C-index= concordance index; se= standard error, DGM= deep grey matter; GM= grey matter; ICA= independent component analysis; 9HPT= 9 hole peg test; SDMT = symbol digit modalities test

**Figure 3.**
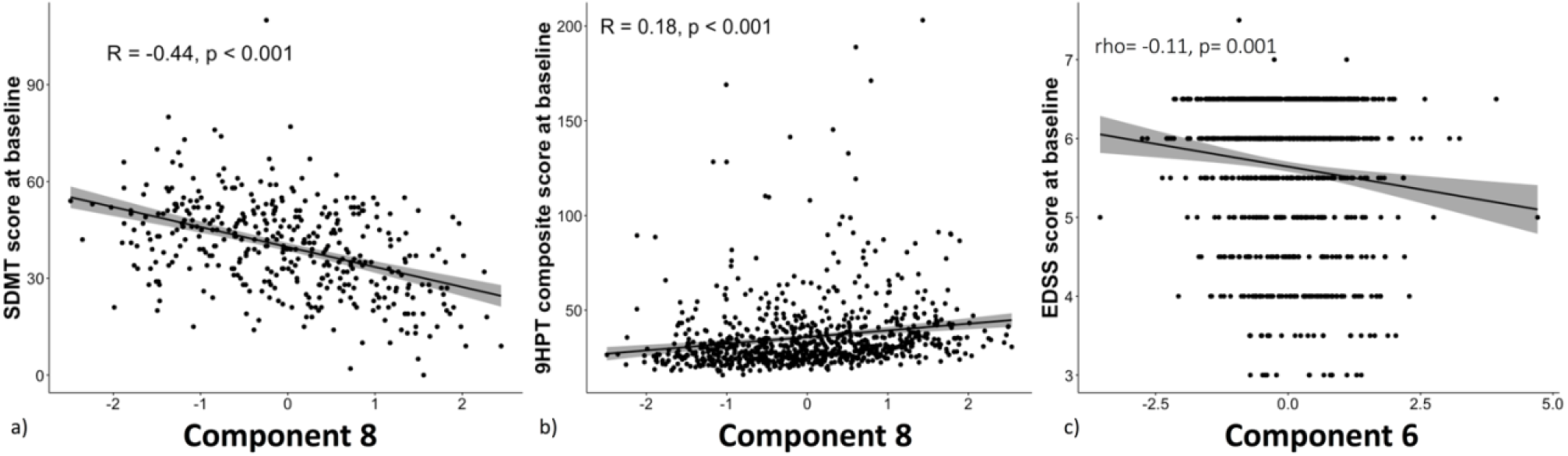
Correlations between baseline ICA-components and baseline EDSS, 9HPT, and SDMT. Legend: Among the 15 stable ICA-component, baseline SDMT score was more strongly associated with a mainly basal ganglia component (component 8). Among the three clinical tests, **(a)** SDMT had the highest correlations with ICA-networks (mainly with component 8). **(b)** 9HPT was associated to the factor loading of component 8. 9HPT and SDMT correlated better with some ICA-networks rather than with any other regional or whole brain MRI measure. **(c)** Among all the 15 networks, component 6 (i.e., cerebellum, brainstem, pons) had the highest correlation with EDSS. We used the false discovery rate (FDR, α = 0.05) to correct for multiple comparisons. Confidence interval band is added to the figure. Acronyms: SDMT= symbol digit modalities test; 9HPT= 9-hole peg test; EDSS= expanded disability status scale

### Stepwise regression and mixed-effect models for cross-sectional analysis

EDSS variability was explained by a model comprising 9 ICA-components, together with whole brain GM and age (F-statistic (818) = 4.91, *p<* 0.001, R^2^ = 0.06, R^2^ corrected = 0.05). The 9 networks encompassed mainly the cerebellum, caudate, putamen, thalamus, precuneus, frontal, parietal, temporal and occipital brain regions (ICA-networks 6, 7, 8, 9, 11, 12, 15, 18, 20, whole brain GM, and age). EDSS was significantly associated with component 6 (β= −0.19, se= 0.07, t(817.87)= −2.76, p <0.01), component 8 (β= 0.17, se= 0.07, t(809.09)= 2.37, p <0.05), component 11 (β= 0.15, se= 0.07, t(776.81)= 2.10, p <0.05), component 20 (β= 0.15, se= 0.07, t(815.87)= 2.08, p <0.05), and whole brain GM (β= −0.17, se= 0.08, t(817.51)= −2.10, p <0.05) (***Table 4*** *in Supplementary materials*).

The best explanatory model for 9HPT comprised ICA-networks 1, 6, 8 (mainly in the superior and middle frontal, cerebellar, basal ganglia, and middle frontal GM), together with whole brain and DGM, age, sex and trial arm (F-statistic(821)= 7.79, *p<* 0.001, R^2^ = 0.07, R^2^ corrected = 0.06). 9HPT was significantly associated with loading of component 6 (β= −2.73, se= 0.71, t(795.74)= −3.87, p <0.001), component 8 (β= 3.11, se= 0.76, t(815.89)= 4.08, p <0.001), whole brain GM volume (β= −2.65, se= 1.32, t(760.01)= −2.00, p <0.05), and age (β= −0.24, se= 0.09, t(793.06)= −-2.56, p <0.01) (***Table 5*** *in Supplementary materials*).

SDMT variability was best explained by a model comprising 7 ICA-components, whole brain GM, lesion load and sex (F-statistic (378) = 17.09, *p<* 0.001, R^2^ = 0.31, R^2^ corrected = 0.29). SDMT was best associated with: component 5 (β= 1.50, se= 0.66, t(349.68)= −2.27, p <0.02), component 7 (β= −1.37, se= 0.60, t(376.56)= −2.29, p <0.02), component 8 (β= −3.61, se= 0.86, t(373.19)= −4.22, p <0.001), component 15 (β=-1.31, se= 0.62, t(374.85)= −2.11, p <0.05), whole brain GM volume (β= 3.95, se= 0.96, t(377.58)= 4.11, p <0.001), lesion load (β= −2.13, se= 0.79, t(373.12)= −2.70, p <0.01), and sex (β= −5.11, se= 1.62, t(374.21)= −-3.16, p <0.005) (***Table 6*** *in Supplementary materials*). According to a multivariable stepwise regression analysis performed on two fitted models (one with the 15 ICA-components and the other with GM, DGM volumes, and lesion load), the first one had a lower Akaike’s Information Criteria (AIC) and higher adjusted R^2^ (respectively, R^2^= 0.27, R^2^-adjusted = 0.26, AIC= 3064.816, p< 0.001; R^2^= 0.23, R^2^-adjusted = 0.22, AIC= 3075.921, p< 0.001), which means that it was better at explaining SDMT variance.

## Predicting disability progression with survival modelling

### Predicting the risk of 12-week confirmed EDSS progression

Data were available for 840 participants (317 males, 523 females, 419 patients under DMT, 421 patients in the placebo group). A total of 28.5% of subjects had 12-week confirmed EDSS progression (***Figure 1*** *in Supplementary Materials)*. None of ICA networks predicted EDSS progression. Baseline caudate volume was the only measure that predicted the EDSS progression (HR= 0.81, 95%CI [0.68: 0.98], p= 0.03) (***Table 7*** *in Supplementary Materials*).

### GM networks predicted 9HPT worsening

Data for 361 subjects were available (134 males, 227 females, 191 patients under DMT, 170 patients in the placebo group). By the last available visit, 42% of participants experienced a worsening in the 9HPT after a mean-time-to-conversion of 1.76 years (***Figure 2*** *in Supplementary materials*). Component 2 (HR= 1.30, 95% CI [1.06:1.60], p<0.01), component 20 (HR= 1.21, 95% CI [1.01:1.45], p<0.05), and DGM (HR= 0.72, 95% CI [0.52:0.99], p= 0.05) predicted the worsening of the 9HPT (***Figure 4*** *and* ***Table 7*** *in Supplementary Materials)*.

**Figure 4.**
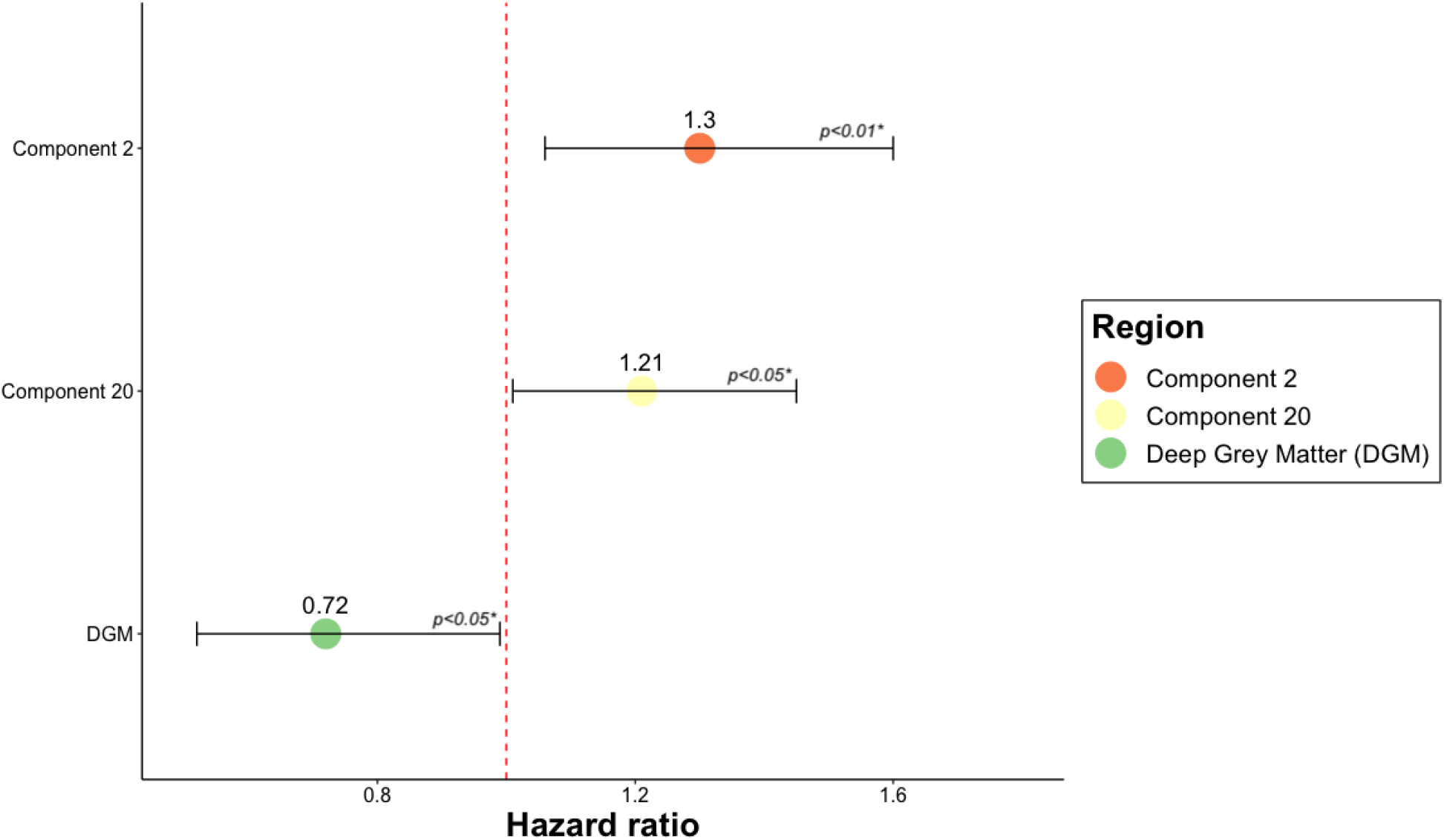
Cox regression models predictive of 9HPT worsening. Figure legend: Hazard ratio (HR) of the statistically significant predictors of 9HPT worsening. The figure shows that 2 GM networks and the volume of the DGM can predict the 9HPT progression. HR higher than 1 indicates that for each standard deviation increase in the corresponding variable there is a higher risk of developing the event. HR values lower than 1 indicates that for each standard deviation decrease in the corresponding variable, there is a higher risk of progressing on 9HPT. Error bars represent the confidence interval. p-values lower that 0.05 represent a statistically significant relative risk of developing a 9HPT progression comparing subjects for each independent variable shown on the vertical axis. Acronyms: DGM= Deep Grey Matter; Component 2 encompasses the temporal lobe, middle cingulate gyrus, precentral gyrus medial segment, posterior cingulate gyrus, parietal lobule, inferior and middle temporal gyrus, parahippocampal gyrus, fusiform gyrus, and entorhinal area. Component 20 consisted of precuneus, posterior cingulate gyrus, middle and superior frontal gyrus, angular gyrus, superior occipital and superior parietal lobule.

### GM networks predicted SDMT worsening

SDMT was available for 360 (140 males, 220 females; 185 under DMT, 175 in the placebo group) subjects. By the last available visit 51% of participants had a 10% worsening [35] in SDMT score after a mean-time-to-conversion of 1.36 years (***Figure 3*** *Supplementary Materials*). SDMT worsening could be predicted by six of ICA-components (component 7, component 8, component 13, component 15, component 17, component 18), lesion load, and thalamus (***Figure 5*** and ***Table 7*** in Supplementary Materials).

**Figure 5.**
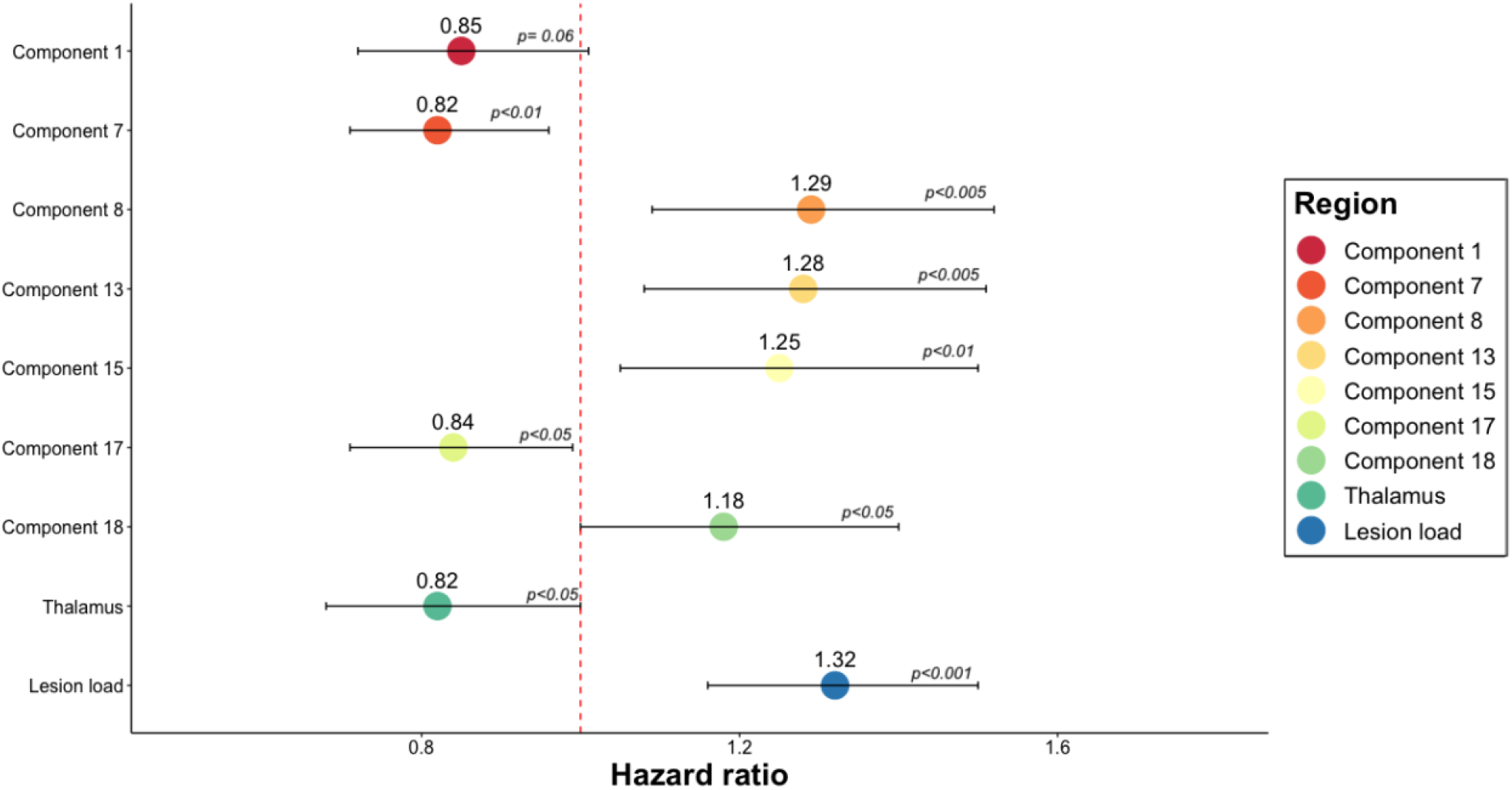
Cox regression models predictive of SDMT worsening. Figure legend. Hazard Ratio (HR) of the statistically significant predictors of SDMT worsening in separate Cox regression models. The figure shows that 6 ICA-components, lesion load, and the volumes of the thalamus could predict the SDMT progression. HR higher than 1 indicates that for each standard deviation increase in the corresponding variable there is a higher risk of developing the event. HR values lower than 1 indicates that for each standard deviation decrease in the corresponding variable, there is a higher risk of progressing on SDMT. For each standard deviation increase in component 8 (encompassing mainly basal ganglia regions), which is inversely related to GM volumes, there was a 29% higher risk of developing SDMT progression. For each standard deviation decrease in the volume of the thalamus there is a 18% increased risk of worsening in SDMT. Error bars represent the confidence interval of HR. p-values lower than 0.05 represent a statistically significant relative risk of developing a SDMT progression for each independent variable shown on the vertical axis. Acronyms: HR= hazard ratio; SDMT= Symbol digit modalities test

### GM networks add value to conventional MRI measures: *Cox-proportional regression analysis*

Models with ICA-components had a higher concordance index (C-index= 0.69, se= 0.025) with respect to models including just conventional MRI measures (C-index= 0.65, se= 0.025) (***Table 3***).

The highest concordance index belonged to a model with all ICA-components (C-index= 0.72, se= 0.021). When compared to models including just conventional MRI measures (C-index= 0.69, se= 0.022), models that include also ICA-components had higher concordance index (C-index > 0.71, se= 0.021) (***Table 3***).

## Discussion

Our main finding is that data-driven network-based measures of GM atrophy can predict physical and cognitive disability in a large cohort of people with SPMS. Further, GM ICA measures correlated more closely with concurrent disability than regional or whole brain GM atrophy, and network-derived measures better predicted disability progression: two of these networks predicted the 9HPT worsening and six networks predicted cognitive disability better than any other assessed MRI measures. Post-hoc analysis showed that network-based measures added value to conventional MRI measures. Interestingly, the network-derived components that correlated with concurrent disability were not necessarily the same as those associated with progression.

We found that ICA-components correlated more closely with disability than regional and whole brain measures. For each disability measure, different ICA-components dominated, encompassing both cortical and subcortical areas. For EDSS it was component 6, which included regions in the cerebellum, brainstem, pons, lingual gyrus, fusiform gyrus, temporal and parietal lobe. For 9HPT components 6 and 8 (thalamus, brainstem, pons, ventral diencephalon, insula, accumbens, caudate, putamen, pallidum, frontal and temporal lobe) were significant, and for the cognitive dysfunction (SDMT) components 1 (superior and middle frontal gyrus, anterior cingulate gyrus), 8, 11 (cuneus, middle and inferior temporal gyrus, occipital pole, calcarine cortex, supramarginal gyrus, superior parietal lobule), 18 (post and precentral gyrus, parahippocampal gyrus, frontal, occipital, parietal lobe; inferior, middle and superior temporal gyrus, supramarginal and fusiform gyrus), and 20 (precuneus, posterior cingulate gyrus, superior and middle frontal gyrus, superior parietal, angular gyrus, superior occipital gyrus) dominated. Although EDSS at baseline was more strongly associated with component 6 than DGM, whole GM atrophy and lesion load measures, the volume of the thalamus in isolation had a higher correlation with EDSS. While whole brain GM and DGM measures span the whole brain, considering several regions not associated with the lower limb functions, component 6 comprised primarily (but not only) areas related to motor functions. Nonetheless, the involvement in this network of brain regions not related to motor functions might have decrease the strength of the association (coefficient of correlation) with EDSS score when compared to the volume of the thalamus taken in isolation. Thalamus is a neuralgic site for motor control, which has already been reported to be associated with EDSS [37].

While the ICA components were identified without prior knowledge of functionally relevant brain regions, their correlations with disability reinforce their usage in predictive models. These components include regions linked with specific neurological and cognitive functions and also those that were both functionally and structurally related. Component 8, which was mainly a basal ganglia-fronto-temporal network, correlated with 9HPT and SDMT at baseline. It includes regions of DGM (thalamus, brainstem, pons, ventral diencephalon, accumbens, caudate, putamen, pallidum) and cortical areas (frontal and temporal lobe), that are known to be involved in motor control, in memory and learning [38]. These regions are also part of the cortico-thalamic, cortico-basal ganglia-thalamo-cortical, and thalamo-cortical pathways that control both sensory and motor information coming from and going to the cortex [39]. Basal ganglia represent a series of interconnected subcortical nuclei (among which the putamen, caudate, accumbens) which are known to be involved in selecting and implementing purposeful actions, facilitating voluntary movements and inhibiting the competing or interfering ones, and controlling non-motor behaviours (e.g., language, working memory, procedural learning, decision making, higher-order process of movement initiation) [39,40]. Moreover, atrophy in the cortex, caudate, putamen, globus pallidus, thalamus and nucleus accumbens have been reported to be associated with lower performance in the SDMT [41]. Component 6 was mainly a cerebellar network. It encompassed brain regions in close proximity and functionally related (cerebellum, brainstem, pons, parietal lobe), already known to be involved in motor functions. It also encompassed the fascicular gyrus and lingual gyrus, which may be functionally related by indirect connections.

Consistent with previous work in predominantly RRMS populations, we found co-varying and clinically relevant patterns of GM atrophy. Previous studies using ICA have identified eight [14] and ten [5] GM components. In the present study, we looked for 20 components, a practical maximum given available computational power, but found 15 could be consistently identified. Our ICA-components only partially overlapped with previously reported GM networks. For instance, component 5 resembled pattern 2 in Steenwijk et al. (2016) study (they both include the middle temporal gyrus, superior temporal gyrus, supramarginal gyrus, postcentral gyrus, and parietal lobule). However, in addition our ICA-component includes other brain regions (cuneus and frontal gyrus) not reported by the previous study. Pattern 8 reported by Bergsland and colleagues (2018) encompasses similar brain areas as in the ICA-component 7 presented here (e.g., calcarine cortex, precuneus, occipital and frontal lobe). However, a perfect match is never present and overall GM networks presented in this study encompass a higher number of brain regions when compared to the above-mentioned studies. Most patterns identified in other studies were not replicated here, nor between studies. There are several potential reasons for this. First, cohort difference: when compared to Bergsland and colleagues’ study (2018), differences in our results may be related to more severe atrophy in SPMS compared to RRMS. Then there are methodological differences, for example, we used GM volumes as input to the ICA instead of cortical thickness, and we allowed for more components to be extracted. Further work is required to resolve these inconsistencies, but a clear overarching finding is that ICA-based analyses identify overlapping components which could otherwise be lost in whole brain and regional atrophy measures, and that these components are clinically relevant.

Our results reinforce the view that GM atrophy in MS represents an interplay between multiple factors, some promoting atrophy and others relatively mitigating it, hence the multiple overlapping ICA components. Several mechanisms may underlie neurodegeneration, including tract mediated anterograde and retrograde degeneration of highly interconnected regions, network-mediated neurodegeneration, meningeal inflammation, mitochondrial failure, hypoxia, and iron deposition [12,42], and these may all occur in the same cortical region or DGM structure. For instance, DGM (in particular the putamen and caudate), is known to present several connections with motor and associative cortices, but appear to be susceptible sites for extensive demyelination and iron deposition [43]. Other brain regions involved in ICA-components are known to be more susceptible to neurodegeneration due to CSF exposure (deep sulci in the temporal pole) and hypoxia (pallidum, precuneus and posterior cingulate). For example, the precuneus and posterior cingulate present extensive connections with several other brain regions and are part of the default brain functional system, known to present under normal condition the highest level of energy consumption [44]. Because neurons require a higher amount of energy to adapt to demyelination [45], this could make highly connected brain regions more susceptible to neurodegeneration. Therefore, several mechanisms can cause the observed patterns of volume changes. Future work with longitudinal ICA studies will investigate this further.

We found baseline ICA components correlated with baseline and longitudinal 9HPT and SDMT measures, and baseline EDSS, but did not predict EDSS progression. While ICA components may have greater relative clinical effects earlier and later in the course of MS, the limitations of disability measures, which are well-recognised for EDSS, might play a role. The EDSS was designed as a composite score, but is heavily weighted toward walking impairment, particularly affecting mid to higher score ranges [46]. In contrast the 9HPT and SDMT were designed as more specific measures and are more likely to reflect the effects of pathology.

Our study has some limitations. In this study we re-analysed data from a large, negative, multicentre study, where MRI data were acquired with different scanners. However, to take into account the effect of centre had on the association and predictive ability of MRI measures on clinical outputs, we used the centre as covariate in our regression models. Non-isotropic 2D T1w scans were acquired, and while we were still able to measure cortical volumes, and identify multiple ICA-components, isotropic 3D scans may enable future studies to identify additional components. We preferred GM volumes over cortical thickness measures as input for ICA analysis because we were also interested in changes of sub-cortical brain regions. Similar to previous works, we smoothed the probability maps to account for inter-subject variability, but this will have reduced sensitivity to small regional effects, albeit offset by the large size of the cohort.

This study focused on GM networks, however MS is a generalised disorder, and so while our ICA components often complemented whole or regional brain GM measures, future work will determine whether white matter regions could increase the predictive accuracy of network-based measures. Because we used data from a phase 3 clinical trial, no data for healthy controls were available thus we cannot exclude whether the same networks would be identified and how they would differ among healthy controls. Regardless of this limitation, network measures had prognostic utility in our cohort.

In conclusion, we have shown that ICA identifies multiple regional patterns of GM atrophy, several of which are relevant to concurrent disability and some predict future progression. Importantly, several of the ICA-derived GM networks were more closely linked with disability, and better able to predict disability progression, than MRI measures currently used as clinical trial outcomes. As the source data for this study was a phase 3 clinical trial, the ICA analysis pipeline we have developed can readily be deployed in future clinical studies. Given the ability of some components to predict future progression, they could be used to stratify those who are more likely to progress.

## Acknowledgments

This study was supported by the International Progressive MS Alliance (IPMSA, award reference number PA-1603-08175). We are grateful to all the IPMSA investigators who have contributed trial data to this study as part of EPITOME: Enhancing Power of Intervention Trials Through Optimized MRI Endpoints network. DC, FB and OC are supported by the NIHR biomedical research centre at UCLH

## Disclosures

The authors have no competing interesting with respect to this research. The full disclosure statement is as follows:

CT has received an ECTRIMS Post-doctoral Research Fellowship in 2015. She has been a consultant for Roche in the last 12 months. DLA has received research grant funding and/ or personal compensation for consulting from Acorda, Adelphi, Alkermes, Biogen, Celgene, Frequency Therapeutics, Genentech, Genzyme, Hoffman-La Roche, Immuene Tolerance Network, Immunotec, MedDay, EMD Serono, Novartis, Pfizer, Receptos, Roche, Sanofi-Aventis, Canadian Institutes of Health Research, MS Society of Canada, and International Progressive MS Alliance; and holds an equity interest in NeuroRx Research. F.B has received compensation for consulting services and/or speaking activities from Bayer Schering Pharma, Biogen Idec, Merck Serono, Novartis, Genzyme, Synthon BV, Roche, Teva, Jansen research and IXICO and is supported by the NIHR Biomedical Research Centre at UCLH. OC has received research grants from the MS Society of Great Britain &Northern Ireland, National Institute for Health Research (NIHR) University College London Hospitals Biomedical Research Centre, EUH2020, Spinal Cord Research Foundation, and Rosetrees Trust. She serves as a consultant for Novartis, Teva, and Roche and has received an honorarium from the American Academy of Neurology as Associate Editor of Neurology and serves on the Editorial Board of Multiple Sclerosis Journal. In the last 3 years DC is a consultant for Biogen and Hoffmann-La Roche. He has received research funding from the International Progressive MS Alliance, the MS Society, and the National Institute for Health Research (NIHR) University College London Hospitals (UCLH) Biomedical Research Centre. AE serves on the Editorial Board of Neurology. He has received speaker’s honoraria from Biogen and At The Limits educational programme, travel support from the National Multiple Sclerosis Society and honorarium from the Journal of Neurology, Neurosurgery and Psychiatry for Editorial Commentaries. AE and FB have equity stake in Queen Square Analytics. EC and JS have nothing to disclose.

### Abbreviations

9HPT: 9-Hole Peg Test
C-index: Concordance Index
CDP: Confirmed Disability Progression
CNS: Central Nervous System
CSF: Cerebro-Spinal Fluid
DGM: Deep Grey Matter
EDSS: Expanded Disability Status Scale
FWHM: ull Width At Half Maximum
GM: Grey matter
HR: Hazard Ratio
ICA: Independent Component Analysis
MRI: Magnetic Resonance Imaging
MS: Multiple Sclerosis
SDMT: Symbol Digit Modalities Test
SPMS: Secondary Progressive Multiple Sclerosis
WM: White Matter

## Notes

### Competing Interest Statement

The authors have declared no competing interest.

### Clinical Trial

NCT01416181

### Author Declarations

The Institutional Review Board at the Montreal Neurological Institute (MNI), Quebec, Canada approved this study under the auspices of International Progressive MS Alliance (Reference number: IRB00010120).

