## Supplementary material for "Predicting disability progression and cognitive worsening in multiple sclerosis using grey matter network measures": Colato_supplementary.pdf

### Image acquisition

A representative MRI protocol from one centre included the following image acquisitions:

(1) 3D T1-weighted: TR= 30 milliseconds, TE= 11.0 milliseconds, slice thickness= 3.0mm, imaging frequency= 63.43, acquisition matrix= 0x256x192x0, flip angle= 30.0, number of phase encoding steps= 192, FoV= 187*250mm;

(2) 2D fast fluid- attenuated inversion recovery (2D FLAIR): with voxel size= 0.98 × 0.98,

TR= 2980.0 milliseconds, TE = 12 milliseconds, slice thickness= 3.0mm, imaging frequency= 63.643, number of phase encoding steps= 258, matrix size = 0x256x256x0, flip angle= 180.0, FoV= 250*250mm;

(3) 2D T2-weighted: with voxel size= 0.98 × 0.98, TR= 5750.0 milliseconds, TE = 86 milliseconds, slice thickness = 3.0mm, imaging frequency= 63.643, number of phase encoding steps= 259, matrix size = 0x256x256x0, flip angle= 180.0, FoV= 250*250mm;

(4) 2D proton density (PD): with voxel size= 0.98 × 0.98, TR= 2980.0 milliseconds, TE = 12 milliseconds, slice thickness = 3.0mm, imaging frequency= 63.643, number of phase encoding steps= 258, matrix size = 0x256x256x0, flip angle= 180.0, FoV= 250*250mm;

### Study-specific template construction

We randomly selected 39 subjects to create a study-specific template in ANTs. The number of subject was chosen to be as large as feasible, based on computation time, and as significantly larger than previous cohorts used for this purpose (e.g., N= 19 (Eshaghi et al., 2014) , N= 20 (Whitwell *et al.*, 2007)). We interpolated the T1w lesion filled scans to 1x1x1mm, and registered them using a rigid body transformation to the MNI152 space (http://www.bic.mni.mcgill.ca/ServicesAtlases/ICBM152NLin2009).

### Whole brain mask

We created a whole brain mask with a twofold purpose: (1) to constrain the ICA analysis at the level of the parenchymal brain, and (2) to identify and label brain regions involved in each ICA-component. We used the mayor voting algorithm implemented in antsJointLabelFusion to fuse the 39 parcellation maps of subjects that had contributed to the template and obtain a single parcellation map where the likelihood that each region corresponds to the labelled one is higher (H. Wang et al., 2013; H. Wang & Yushkevich, 2013). We used this atlas-mask to label brain regions involved in each component. We have then thresholded out not brain structures (e.g., ventricles, meninges, etc.), binarised the output and used the obtained mask to mask the processed GM probability maps and reduce the computational requirements of the ICA analysis.

### Supplementary Tables

***Supplementary Table 1.*** Results of the spatial cross-correlations between the 20 components obtained in each of the four sub-sample and from the entire cohort

| Network | Sub-sample 1 vs. 2 | Sub-sample 1 vs. 3 | Sub-sample 1 vs.4 | Entire cohort vs. sub-sample 1 | Sub-sample 2 vs. 3 | Sub-sample 2 vs. 4 | Entire cohort vs. sub-sample 2 | Sub-sample 3 vs. 4 | Entire cohort vs. sub-sample 3 | Entire cohort vs. sub-sample 4 |
|---|---|---|---|---|---|---|---|---|---|---|
| **1** | 0.86 | 0.80 | 0.84 | 0.89 | 0.82 | 0.88 | 0.91 | 0.82 | 0.84 | 0.88 |
| **2** | 0.88 | 0.88 | 0.87 | 0.90 | 0.92 | 0.90 | 0.95 | 0.89 | 0.93 | 0.92 |
| **3** | 0.41 | 0.30 | 0.52 | 0.35 | 0.66 | 0.36 | 0.54 | 0.48 | 0.63 | 0.73 |
| **5** | 0.37 | 0.61 | 0.50 | 0.43 | 0.28 | 0.16 | 0.41 | 0.40 | 0.42 | 0.14 |
| **6** | 0.94 | 0.94 | 0.93 | 0.94 | 0.95 | 0.95 | 0.97 | 0.94 | 0.95 | 0.96 |
| **7** | 0.10 | 0.27 | 0.20 | 0.22 | 0.55 | 0.51 | 0.63 | 0.60 | 0.73 | 0.75 |
| **8** | 0.93 | 0.90 | 0.88 | 0.95 | 0.88 | 0.93 | 0.95 | 0.82 | 0.90 | 0.91 |
| **9** | 0.62 | 0.61 | 0.64 | 0.75 | 0.56 | 0.69 | 0.77 | 0.62 | 0.72 | 0.76 |
| **11** | 0.78 | 0.62 | 0.41 | 0.78 | 0.53 | 0.35 | 0.74 | 0.53 | 0.81 | 0.52 |
| **12** | 0.48 | 0.69 | 0.47 | 0.82 | 0.51 | 0.20 | 0.55 | 0.24 | 0.83 | 0.39 |
| **13** | 0.85 | 0.78 | 0.86 | 0.92 | 0.76 | 0.83 | 0.89 | 0.77 | 0.82 | 0.90 |
| **15** | 0.41 | 0.42 | 0.52 | 0.52 | 0.20 | 0.33 | 0.18 | 0.43 | 0.69 | 0.59 |
| **17** | 0.40 | 0.63 | 0.60 | 0.69 | 0.52 | 0.46 | 0.51 | 0.66 | 0.81 | 0.71 |
| **18** | 0.63 | 0.51 | 0.44 | 0.53 | 0.55 | 0.61 | 0.61 | 0.33 | 0.82 | 0.36 |
| **20** | 0.69 | 0.67 | 0.62 | 0.72 | 0.80 | 0.70 | 0.81 | 0.75 | 0.90 | 0.80 |

Table legend: The table reports the spatial cross-correlations results for the stable ICA-components (i.e., showed a statistically significant correlations ($p<0.05$) in all 4 sub-folds and in the entire cohort).

***Supplementary Table 2.*** Correlations between the loading values of ICA-components and whole brain GM volumes, SDMT, EDSS, and 9HPT score

| GM NETWORKS | WHOLE BRAIN GM VOLUME | EDSS N= 830 | 9HPT N= 829 | SDMT N= 391 |
|---|---|---|---|---|
| Component 1 | r= 0.08<br>95% CI<br>[0.01:0.14]<br>p<0.05* | rho= - 0.01<br>p= 0.79 | r= - 0.09<br>95% CI<br>[- 0.15: -0.02]<br>p= 0.07 | r= 0.22<br>95% CI<br>[0.13:0.32]<br>p<0.001* |
| Component 2 | r= - 0.25<br>95%CI<br>[-0.3:-0.19]<br>p<0.001* | rho= 0.08<br>p= 0.15 | r= 0.02<br>95% CI<br>[-0.05: 0.09]<br>p= 1 | r= - 0.07<br>95% CI<br>[- 0.17:0.03]<br>p= 0.25 |
| Component 3 | r= 0.18<br>95%CI<br>[0.11:0.23]<br>p<0.001* | rho= - 0.05<br>p= 0.22 | r= - 0.014<br>95% CI<br>[-0.08:0.05]<br>p= 1 | r= 0.04<br>95% CI<br>[- 0.06:0.14]<br>p= 0.46 |
| Component 5 | r= - 0.16<br>95%CI<br>[-0.22:-0.09]<br>p<0.001* | rho= - 0.03<br>p= 0.39 | r= - 0.04<br>95% CI<br>[- 0.1: 0.03]<br>p= 0.79 | r= 0.07<br>95% CI<br>[- 0.03:0.16]<br>p= 0.26 |
| Component 6 | r= - 0.09<br>95% CI<br>[-0.15:-0.02]<br>p<0.001* | rho= - 0.11<br>p= 0.02* | r= - 0.15<br>95% CI<br>[-0.21: -0.08]<br>p< 0.001* | r= 0.10<br>95% CI<br>[0:0.19]<br>p= 0.10 |
| Component 7 | r= - 0.07<br>95% CI<br>[-0.13:-0.01]<br>p= 0.05* | rho= 0.04<br>p= 0.36 | r= - 0.01<br>95% CI<br>[- 0.08: 0.06]<br>p= 1 | r= - 0.06<br>95% CI<br>[- 0.16:0.04]<br>p= 0.26 |
| Component 8 | r= - 0.18<br>95% CI<br>[-0.24:-0.12]<br>p<0.001* | rho= 0.06<br>p= 0.22 | r= 0.18<br>95% CI<br>[0.11:0.24]<br>p< 0.001* | r= - 0.44<br>95% CI<br>[- 0.52: - 0.36]<br>p<0.001* |

| | | | | |
|---|---|---|---|---|
| Component 9 | r= 0.20<br>95% CI<br>[0.14:0.26]<br>p<0.001* | rho= 0.06<br>p= 0.21 | r= 0.07<br>95% CI<br>[0.0:1.14]<br>p= 0.19 | r= - 0.01<br>95% CI<br>[- 0.11:0.09]<br>p= 0.85 |
| Component 11 | r= - 0.12<br>95% CI<br>[-0.17:-0.05]<br>p<0.001* | rho= 0.07<br>p= 0.12 | r= - 0.05<br>95% CI<br>[- 0.12: 0.02]<br>p= 0.43 | r= 0.16<br>95% CI<br>[0.06:0.25]<br>p<0.01* |
| Component 12 | r= - 0.17<br>95% CI<br>[-0.23:-0.11]<br>p<0.001* | rho= - 0.04<br>p= 0.35 | r= 0.01<br>95% CI<br>[- 0.06:0.08]<br>p= 1 | r= - 0.12<br>95% CI<br>[- 0.21: -0.02]<br>p<0.05* |
| Component 13 | r= - 0.38<br>95% CI<br>[-0.43:-0.33]<br>p<0.001* | rho= 0.07<br>p= 0.16 | r= 0.03<br>95% CI<br>[- 0.04:0.1]<br>p= 0.92 | r= - 0.12<br>95% CI<br>[- 0.21: - 0.02]<br>p<0.05* |
| Component 15 | r= - 0.01<br>95% CI<br>[-0.07:0.06]<br>p= 0.87 | rho= 0.07<br>p= 0.18 | r= -0.01<br>95% CI<br>[- 0.07:0.06]<br>p= 1 | r= - 0.13<br>95% CI<br>[- 0.23: - 0.03]<br>p<0.05* |
| Component 17 | r= 0.19<br>95% CI<br>[0.13:0.25]<br>p<0.001* | rho= 0.02<br>p= 0.65 | r= 0.01<br>95% CI<br>[- 0.06:0.08]<br>p= 1 | r= 0.03<br>95% CI<br>[- 0.07:0.13]<br>p= 0.59 |
| Component 18 | r= - 0.28<br>95% CI<br>[-0.33:-0.22]<br>p<0.001* | rho= 0.08<br>p= 0.19 | r= - 0.002<br>95% CI<br>[- 0.07: 0.07]<br>p= 1 | r= - 0.15<br>95% CI<br>[- 0.25: - 0.05]<br>p<0.01* |
| Component 20 | r= 0.28<br>95% CI<br>[0.22:0.33]<br>p<0.001* | rho= 0.05<br>p= 0.22 | r= 0<br>95% CI<br>[- 0.07-0.07]<br>p= 1 | r= 0.18<br>95% CI<br>[0.08: 0.27]<br>p<0.005* |

Acronyms: GM= grey matter, EDSS= expanded disability status scale; 9HPT= nine hole peg test; SDMT= symbol digit modalities test; CI= confidence interval

*Statistically significant after False Discovery Rate correction

We report the Benjamini-Hochberg Adjusted P values

**Supplementary Table 3.** Correlations between the SDMT, EDSS, and 9HPT score at baseline and the lesion load, DGM, GM, and smaller regional volumes.

| **MRI MEASURES** | **EDSS**<br>**N= 830** | **9HPT**<br>**N= 829** | **SDMT**<br>**N= 391** |
|---|---|---|---|
| Whole brain GM | rho= - 0.09<br>p= 0.052 | r= - 0.01<br>95% CI<br>[- 0.08: 0.06]<br>p= 1 | r= 0.22<br>95% CI<br>[0.12:0.31]<br>p<0.001* |
| Deep GM | rho= - 0.07<br>p<0.05* | r= 0.01<br>95% CI<br>[- 0.06:0.08]<br>p= 0.83 | r= 0.13<br>95% CI<br>[0.03:0.23]<br>p<0.001** |
| Lesion load | rho= 0.07<br>p= 0.06 | r= 0.15<br>95% CI<br>[0.08:0.22]<br>p<0.001* | r= -0.43<br>95% CI<br>[-0.51:-0.35]<br>p<0.001* |
| Thalamus | rho= - 0.12<br>p< 0.005* | r= - 0.13<br>95% CI<br>[- 0.20: - 0.07]<br>p<0.001* | r= 0.41<br>95% CI<br>[0.32:0.49]<br>p<0.001* |
| Precuneus | rho= - 0.05<br>p= 0.13 | r= 0.01<br>95% CI<br>[- 0.06:0.08]<br>p= 0.94 | r= 0.21<br>95% CI<br>[0.11:0.30]<br>p<0.001* |
| Caudate | R<br>ho= - 0.07<br>p= 0.051 | r= - 0.02<br>95% CI<br>[-0.09:0.04]<br>p= 0.79 | r= 0.15<br>95% CI<br>[0.05:0.24]<br>p<0.001* |
| Putamen | rho= -0.07<br>p< 0.05* | r= - 0.05<br>95% CI<br>[- 0.12:0.02]<br>p= 0.27 | r= 0.31<br>95% CI<br>[0.22:0.40]<br>p<0.001* |
| Pallidum | rho= -0.08<br>p< 0.05* | r= -0.01<br>95% CI<br>[- 0.14: - 0.01]<br>p= 0.08 | r= 0.31<br>95% CI<br>[0.22:0.40]<br>p<0.001* |

Acronyms: GM= grey matter, EDSS= expanded disability status scale; 9HPT= nine-hole peg test; SDMT= symbol digit modalities test; CI= confidence interval

*Statistically significant after False Discovery Rate correction

We report the Benjamini-Hochberg Adjusted P values

***Supplementary Table 4.*** Regression analysis for EDSS

| **Predictor** | **Estimate** | **Std.Error** | **df** | **t-value** | **p-value** |
|---|---|---|---|---|---|
| **Component 6** | -0.19 | 0.07 | 817.87 | -2.76 | 0.006 ** |
| **Component 7** | 0.11 | 0.07 | 815.48 | 1.67 | 0.10 |
| **Component 8** | 0.17 | 0.07 | 809.09 | 2.37 | 0.02* |
| **Component 9** | 0.11 | 0.07 | 814.46 | 1.62 | 0.11 |
| **Component 11** | 0.15 | 0.07 | 776.81 | 2.10 | 0.04* |
| **Component 12** | -0.11 | 0.07 | 760.98 | -1.61 | 0.11 |
| **Component 15** | 0.11 | 0.07 | 817.69 | 1.70 | 0.09 |
| **Component 18** | 0.08 | 0.07 | 740.56 | 1.06 | 0.29 |
| **Component 20** | 0.15 | 0.07 | 815.87 | 2.08 | 0.04* |
| **Whole brain GM** | -0.17 | 0.08 | 817.51 | -2.10 | 0.04* |
| **Age** | 0.013 | 0.01 | 818.00 | 1.44 | 0.15 |

Table legend: We used a stepwise regression analysis to identify the best explanatory variables for Expanded Disability Status Scale (EDSS) among the 15 GM networks, the volume of whole brain GM and deep GM, lesion, age, sex, and trial arm. We then fitted a mixed effect model to identify the predictive potential of the selected variables.

Acronyms: GM= grey matter, Std.Error = standard error; df= degrees of freedom

***Supplementary Table 5.*** Regression analysis for 9HPT

| Predictor | Estimate | Std.Error | df | t-value | p-value |
|---|---|---|---|---|---|
| **Component 1** | -1.24 | 0.73 | 678.75 | -1.70 | 0.09 |
| **Component 6** | -2.73 | 0.71 | 795.74 | -3.87 | p<0.001*** |
| **Component 8** | 3.11 | 0.76 | 815.89 | 4.08 | p<0.001*** |
| **Whole brain GM volume** | -2.65 | 1.32 | 760.01 | -2.00 | p<0.05* |
| **Deep GM volume** | 3.51 | 1.81 | 811.07 | 1.94 | 0.053 |
| **Age** | -0.24 | 0.09 | 793.06 | -2.56 | 0.01* |
| **Sex** | 2.94 | 1.76 | 812.947 | 1.67 | 0.10 |
| **Trail Arm** | 1.86 | 1.32 | 736.96 | 1.42 | 0.16 |

Table legend: We used a stepwise regression analysis to identify the best explanatory variables for 9 Hole Peg Test among the 15 GM networks, the volume of whole brain GM and deep GM, lesion, age, sex, and trial arm. We then fitted a mixed effect model to identify the predictive potential of the selected variables.

Acronyms: GM= grey matter, Std.Error = standard error; df= degrees of freedom

***Supplementary Table 6.*** Regression analysis for SDMT

| Predictor | Estimate | Std.Error | df | t-value | p-value |
|---|---|---|---|---|---|
| **Component 5** | 1.50 | 0.66 | 349.68 | 2.27 | 0.02* |
| **Component 6** | 0.88 | 0.68 | 377.10 | 1.29 | 0.20 |
| **Component 7** | -1.37 | 0.60 | 376.56 | -2.29 | 0.02* |
| **Component 8** | -3.61 | 0.86 | 373.19 | -4.22 | 0.001*** |
| **Component 11** | 0.90 | 0.715 | 373.85 | 1.26 | 0.21 |
| **Component 15** | -1.31 | 0.62 | 374.85 | -2.11 | 0.04* |
| **Component 20** | 0.79 | 0.71 | 374.80 | 1.12 | 0.26 |
| **Whole brain GM volume** | 3.95 | 0.96 | 377.58 | 4.11 | 0.001*** |
| **Lesion load** | -2.13 | 0.79 | 373.12 | -2.70 | 0.007** |
| **Sex** | -5.11 | 1.62 | 374.21 | -3.16 | 0.002** |

Table legend: We used a stepwise regression analysis to identify the best explanatory variables for Symbol Digit modalities test (SDMT) among the 15 GM networks, the volume of whole brain GM and deep GM, lesion, age, sex, and trial arm. We then fitted a mixed effect model to identify the predictive potential of the selected variables.

Acronyms: GM= grey matter, Std.Error = standard error; df= degrees of freedom

**Supplementary Table 7.** Survival analysis for the EDSS, 9HPT, and SDMT progression

| **Predictors** | **EDSS progression confirmed at 3 months** | | | **20% 9HPT worsening** | | | **10% SDMT worsening** | | |
|---|---|---|---|---|---|---|---|---|---|
| **Brain regions** | **HR** | **95% CI** | **p-value** | **HR** | **95% CI** | **p-value** | **HR** | **95% CI** | **p-value** |
| **Component 1** | 1.00 | 0.86-1.16 | 0.97 | 1.18 | 0.96-1.45 | 0.11 | 0.85 | 0.72-1.01 | 0.06 |
| **Component 2** | 1.10 | 0.94-1.27 | 0.24 | 1.30 | 1.06-1.60 | 0.01** | 1.03 | 0.86-1.22 | 0.76 |
| **Component 3** | 1.02 | 0.88-1.17 | 0.82 | 1.03 | 0.86-1.25 | 0.73 | 0.97 | 0.82-1.15 | 0.69 |
| **Component 5** | 0.90 | 0.78-1.04 | 0.15 | 0.92 | 0.76-1.11 | 0.38 | 1.07 | 0.91-1.26 | 0.44 |
| **Component 6** | 1.04 | 0.90-1.20 | 0.64 | 0.88 | 0.72-1.08 | 0.21 | 0.97 | 0.81-1.16 | 0.74 |
| **Component 7** | 1.05 | 0.91-1.21 | 0.50 | 0.86 | 0.72-1.03 | 0.10 | 0.82 | 0.71-0.96 | 0.01* |
| **Component 8** | 1.04 | 0.90-1.20 | 0.58 | 1.02 | 0.84-1.24 | 0.84 | 1.29 | 1.09-1.52 | p<0.003*** |
| **Component 9** | 0.98 | 0.84-1.13 | 0.75 | 0.95 | 0.77-1.16 | 0.58 | 0.91 | 0.76-1.09 | 0.30 |
| **Component 11** | 0.98 | 0.85-1.14 | 0.82 | 1.03 | 0.85-1.24 | 0.76 | 0.93 | 0.78-1.12 | 0.46 |
| **Component 12** | 1.01 | 0.88-1.17 | 0.88 | 0.85 | 0.71-1.02 | 0.08 | 1.14 | 0.96-1.34 | 0.13 |
| **Component 13** | 1.01 | 0.87-1.16 | 0.95 | 0.94 | 0.78-1.13 | 0.49 | 1.28 | 1.08-1.51 | p<0.005*** |
| **Component 15** | 0.99 | 0.86-1.14 | 0.87 | 1.07 | 0.89-1.28 | 0.50 | 1.25 | 1.05-1.50 | 0.01** |
| **Component 17** | 0.97 | 0.84-1.12 | 0.68 | 0.89 | 0.74-1.07 | 0.22 | 0.84 | 0.71-0.99 | 0.04* |
| **Component 18** | 1.08 | 0.94-1.25 | 0.30 | 1.14 | 0.95-1.36 | 0.17 | 1.18 | 1.00-1.40 | 0.046* |
| **Component 20** | 1.05 | 0.91-1.21 | 0.55 | 1.21 | 1.01-1.45 | 0.04* | 1.00 | 0.85-1.17 | 0.96 |
| **Whole GM** | 0.98 | 0.61-1.02 | 0.07 | 0.90 | 0.71-1.14 | 0.38 | 0.83 | 0.67-1.03 | 0.09 |
| **DGM** | 0.79 | 0.81-1.19 | 0.86 | 0.72 | 0.52-0.99 | 0.05* | 0.95 | 0.71-1.26 | 0.72 |
| **Lesion load** | 0.99 | 0.87-1.12 | 0.83 | 0.98 | 0.82-1.16 | 0.79 | 1.32 | 1.16-1.50 | p<0.001**** |
| **Thalamus** | 0.96 | 0.81-1.14 | 0.63 | 1.01 | 0.81-1.26 | 0.90 | 0.82 | 0.68-1.00 | 0.048* |
| **Precuneus** | 1.01 | 0.85-1.21 | 0.89 | 1.08 | 0.88-1.33 | 0.47 | 0.98 | 0.81-1.19 | 0.84 |
| **Putamen** | 0.91 | 0.77-1.08 | 0.30 | 0.96 | 0.77-1.20 | 0.74 | 0.85 | 0.70-1.03 | 0.10 |
| **Pallidum** | 0.91 | 0.77-1.09 | 0.32 | 0.99 | 0.80-1.24 | 0.95 | 0.83 | 0.68-1.02 | 0.08 |
| **Caudate** | 0.81 | 0.67-0.97 | 0.03* | 0.93 | 0.74-1.17 | 0.54 | 1.05 | 0.86-1.30 | 0.62 |

*Caption: *p<0.05 ** p<0.01 ***p<0.005 ****p<0.001*

Table legend: We report here the results for the univariate Cox regression models for all the assessed independent variables (predictors). The baseline volume of the caudate predicted the EDSS progression confirmed at 3 months. For each standard deviation unit decrease in the baseline caudate volume the risk of developing a confirmed disability progression increased by the 19%. Two ICA-components (component 2 and component 20), and the baseline volume of the DGM predicted the 20% worsening of the 9HPT. Six ICA-components, lesion load, and the volume of the thalamus predicted the 10% SDMT worsening.

Acronyms: EDSS= expanded disability status scale; 9HPT= nine-hole peg test; SDMT= symbol digit modalities test; HR= hazard ratio, CI= confidence interval; DGM= deep grey matter

*Supplementary Figure 1*. Cumulative hazard function for the EDSS progression confirmed at 3 months.

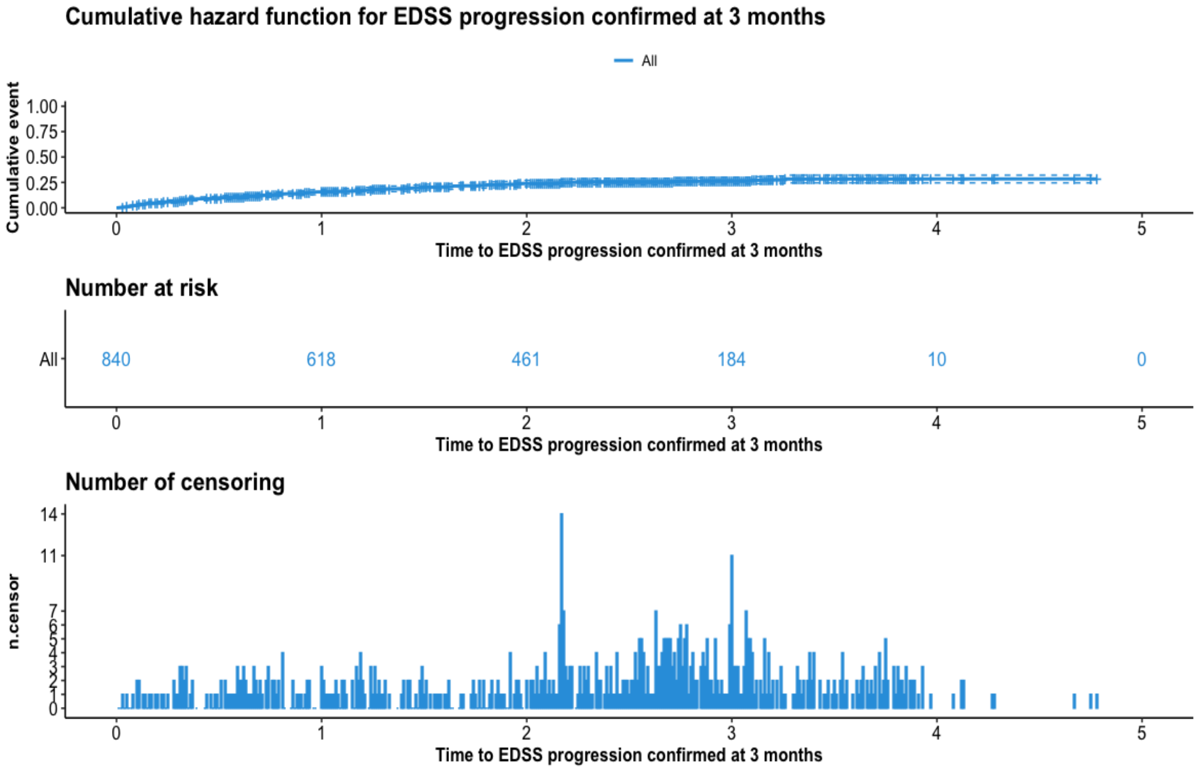


*Figure legend:* The figure shows the cumulative hazard function for the EDSS progression confirmed at 3 months. By the end of the study, the 28.5% of patients reported a 12-week confirmed EDSS progression. The number of censored and the number of subjects at risk of developing the EDSS progression at each timepoint are also reported

*Supplementary Figure 2.* Cumulative hazard function for the 20% 9HPT worsening

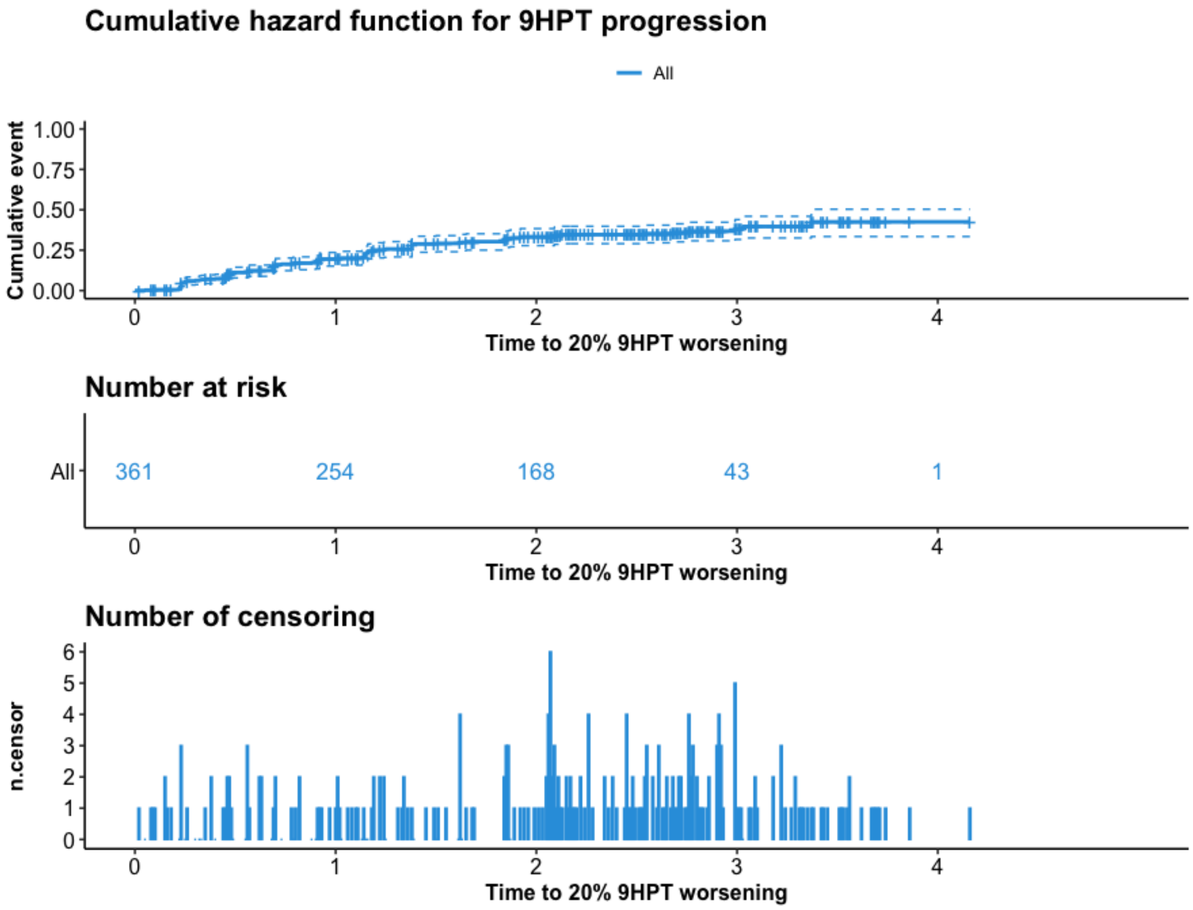


*Figure legend:* The figure shows the cumulative hazard function for the 20% 9HPT worsening. By the end of the study, the 42% of patients reported a 9HPT progression. The number of censored and the number of subjects at risk of developing the 9HPT progression at each timepoint are also reported

*Supplementary Figure 3.* Cumulative hazard function for the 10% SDMT worsening

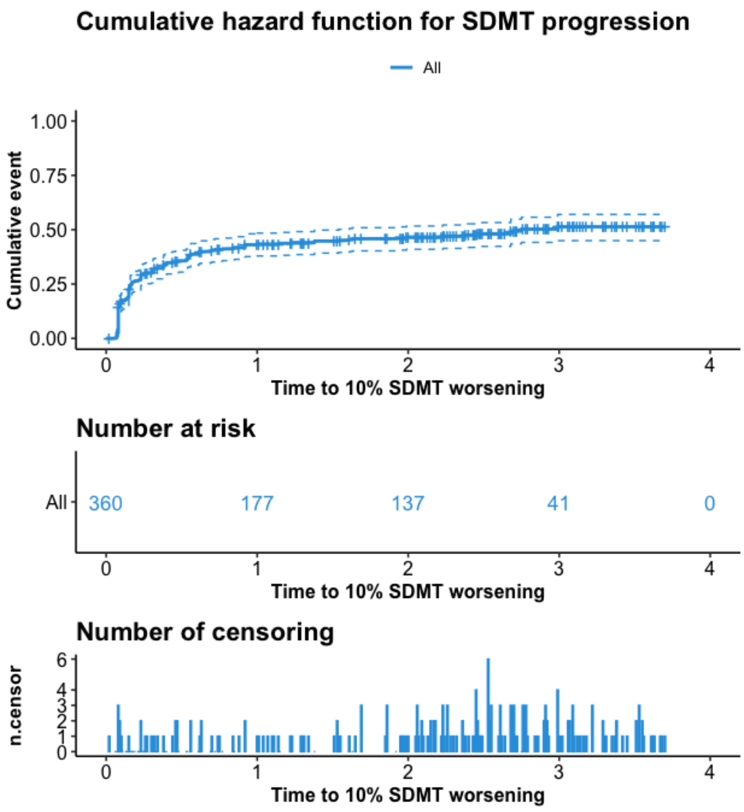


*Figure legend:* The figure shows the cumulative hazard function for the 10% SDMT worsening. By the end of the study, the 51% of subjects reported a SDMT progression. The number of censored and the number of subjects at risk of developing the SDMT progression at each timepoint are also reported
